# Estimate of COVID-19 case prevalence in India based on surveillance data of patients with severe acute respiratory illness

**DOI:** 10.1101/2020.04.14.20065342

**Authors:** Padmanaban Venkatesan

## Abstract

In absence of extensive testing for SARS-CoV-2, true prevalence of COVID-19 cases in India remain unknown. In this study, a conservative estimate of prevalence of COVID-19 is calculated based on the age wise COVID-19 positivity rate among patients with severe respiratory illness as reported by Indian Council of Medical Research. Calculations in the study estimates a cumulative number of 17151 COVID-19 positive cases by the end of April 2, 2020.

## Introduction

The first case of corona virus disease 2019(covid-19) in India due to the severe acute respiratory syndrome coronavirus-2(SARS-CoV-2) was reported on30 ^th^ January 2020 (1). Despite more than 1.5 million international arrivals to India from January 18 to March 23, India had tested only 5900 individuals for SARS-CoV-2 up until 13 ^th^ March, 2020 (2,3). Since then, testing was increased but at total tests of 202551 as of 13 ^th^ April, 2020, India is still among the countries doing least number of tests per 1000 people (4,5). In absence of extensive testing, the incidence of COVID-19 cases could have been vastly underreported. To study the extent of spread of COVID-19 cases, ICMR had tested for SARS-CoV-2 in samples from patients admitted with severe acute respiratory illness (SARI) in multiple centres spread across India from Feb 15 to Apr 2, 2020 (6). In this study I have estimated a conservative number of possible COVID-19 cases based on the data from ICMR on surveillance of patients with SARI.

## Method

Data on age wise and total SARS-CoV-2 positivity rate among SARI patients was extracted from ICMR’s publication of the surveillance (6).

Age wise prevalence of SARI in India during the period of surveillance was estimated as follows. Data on age wise annual number of deaths due to lower respiratory tract infections (LRI) was extracted from Global Burden of Disease Study 2017 (GBD 2017) results, available from http://ghdx.healthdata.org/gbd-results-tool. Assuming that the deaths due to LRI would be a fraction of SARI cases, age wise prevalence of SARI can be estimated with data on case fatality rate of SARI. Since case fatality rate of SARI was not available for India, data on age wise case fatality rate of SARI among African countries was used assuming it would be similar to case fatality rate in India (7). To account for possible seasonality in prevalence of SARI, the resulting number of cases for the period of surveillance was divided by 2 assuming the prevalence of SARI during the months of March and April would have been half the average prevalence.

Total number of possible hospitalized COVID-19 cases during the period of surveillance was estimated age wise from the prevalence of SARI cases calculated as described above and SARS-CoV-2 positivity rate among SARI cases.

Total number of age wise prevalence of COVID-19 cases was estimated from possible hospitalized COVID-19 cases as calculated above and from published data on age wise hospitalisation rate among COVID-19 cases (8).

Calculations were done in R.

## Results

Table 1 shows the total number of estimated COVID-19 cases in India. A total of 8100 cases could have had COVID-19 during the period of March 22 to 28 and another 9051 during the period of March 29 to Apr 2. Thus, by end of Apr 2, the cumulative number of COVID-19 cases must have been at least 17151. The actual cumulative number of laboratory confirmed cases stood at 1024 on March 28 and 2545 on April 2 resulting in an ascertainment ratio of 0.13 and 0.15 respectively.

**Table 1:** Estimation of COVID-19 cases in India

|  | Estimated SARI cases |  | Estimated hospitalised COVID-19 cases |  | Estimated total COVID-19 cases |  |
| --- | --- | --- | --- | --- | --- | --- |
| Age group (years) | Mar 22 to 28 | March 29 to Apr 2 | Mar 22 to 28 | March 29 to Apr 2 | Mar 22 to 28 | March 29 to Apr 2 |
| 0-9 | 16510 | 11793 | 81 | 91 | * | * |
| 10-19 | 1754 | 1253 | 0 | 0 | * | * |
| 20-29 | 1273 | 909 | 7 | 8 | 719 | 803 |
| 30-39 | 1458 | 1041 | 11 | 12 | 323 | 361 |
| 40-49 | 1934 | 1381 | 45 | 51 | 1064 | 1189 |
| 50-59 | 3800 | 2714 | 172 | 192 | 2109 | 2357 |
| 60-69 | 7712 | 5508 | 278 | 311 | 2356 | 2633 |
| 70-79 | 9738 | 6956 | 178 | 199 | 1075 | 1201 |
| >80 | 8239 | 5885 | 83 | 93 | 453 | 506 |

\*Cannot be estimated due to very low hospitalisation rate among this age group

Fig 1 shows plot of estimated number of cases with exponential fit and incidence of laboratory confirmed cases.

**Fig 1:**
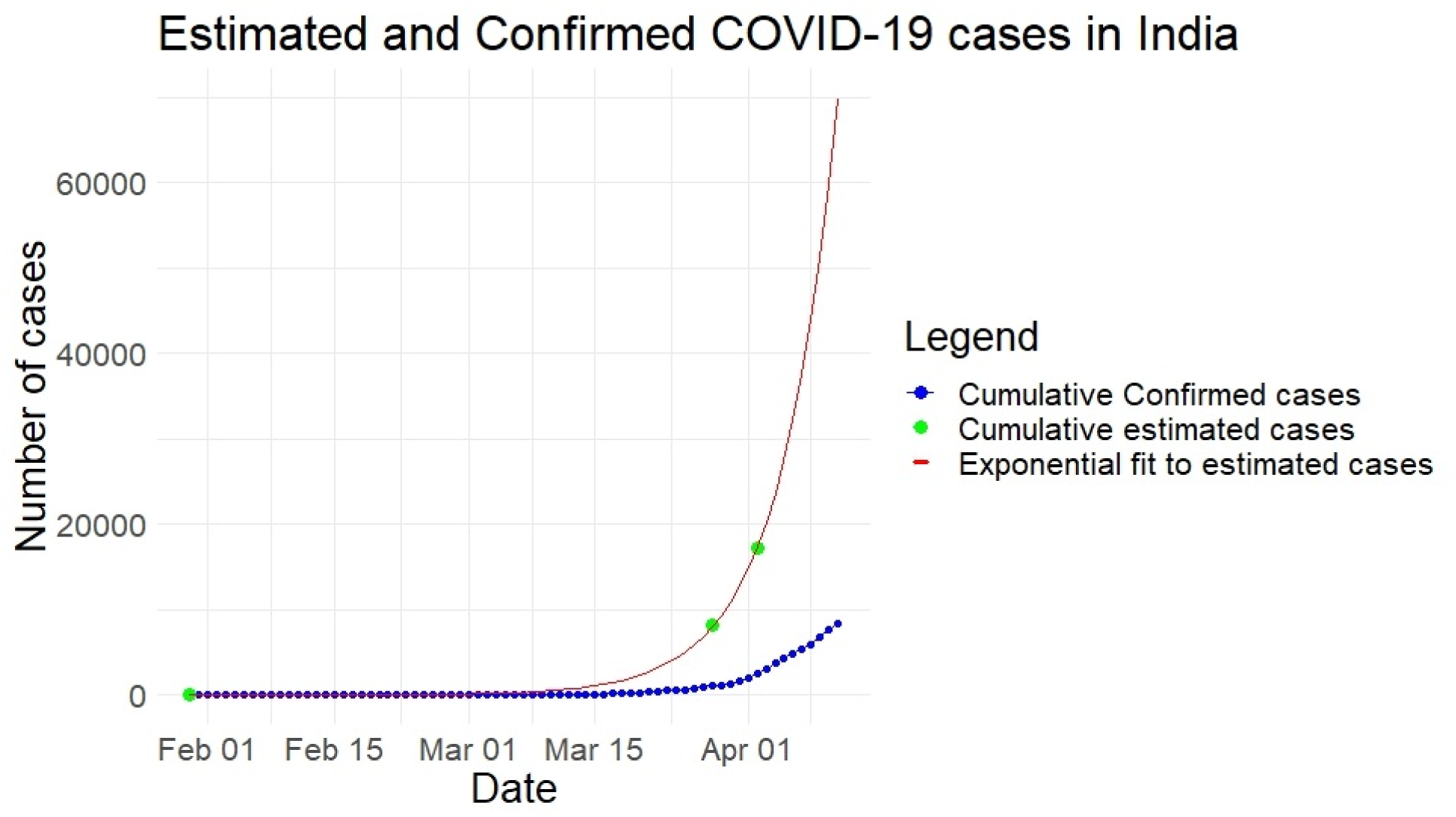
Estimated and confirmed COVID-19 cases in India.

## Conclusion

Conservative estimate of COVID-19 cases in India based on ICMR’s surveillance data of SARI cases show that low testing rates in India could be vastly underestimating the actual number of cases.

## Data Availability

All data used in the manuscript is publicly available

